# Association between polygenic risk and survival in breast cancer patients

**DOI:** 10.1101/2024.10.31.24315531

**Authors:** Danielle E. Kurant, Stefan Groha, Yi Ding, Chris German, Wei Wang, Julie M. Granka, Michael V. Holmes, 23andMe Research Team, Suyash S. Shringarpure, Alexander Sasha Gusev

**Affiliations:** Department of Medical Oncology, Dana-Farber Cancer Institute and Harvard Medical School, Boston, MA 02215; 23andMe, Inc., Sunnyvale, CA 94086

## Abstract

Polygenic risk scores (PRS) estimate an individual’s germline genetic predisposition to a quantitative trait and/or risk of disease. Several PRS have been developed for cancer risk with the goal of improved risk screening. However, little is known about the association with subsequent outcomes for individuals who develop cancer. Here, we sought to establish whether PRS for cancer risk and other common traits may influence survival for patients with cancer. We conducted a PRS survival analysis using 23,770 European cancer patients from the Dana-Farber Cancer Institute Profile cohort. We identified an association between PRS for breast cancer risk and longer patient survival (HR = 0.89, p = 1.50×10^-4^, <5% FDR), implying that individuals at high genetic risk had better outcomes. High PRS individuals were also significantly less likely to harbor somatic TP53 mutations, consistent with having less aggressive tumors. This association persisted when including tumor grade and became more protective when restricting to ER-negative tumors (HR = 0.78, p = 1.69×10^-4^). Potential confounders such as hormone receptor status, age, grade, stage, and ER-targeted therapy did not fully explain this association, nor was there statistical evidence of index event bias at individual variants. We did not observe significant associations between cancer risk and survival for other cancers, suggesting that this mechanism may be largely unique to breast cancer. However, we did observe associations between shorter survival and type 2 diabetes, bipolar, and pancreatitis PRS (1% FDR). These findings suggest that higher germline risk may predispose individuals to less aggressive breast cancer tumors and provide novel insights into breast cancer development and prognosis.

## Introduction

Polygenic risk scores (PRS) summarize a patient’s genetic risk for a variety of phenotypes. PRS may serve as a means of identifying individuals at increased genetic risk for cancer who lack significant family history or other known risk factors. Cancer risk PRS have been shown to predict cancer incidence in independent cohorts. Recently, cancer risk PRS have been associated with specific somatic alterations and molecular subtypes ^1–8^. However, little is known about PRS association with clinical outcomes.

PRS have shown promise in predicting breast cancer risk in addition to the risk of developing contralateral breast cancer ^7, 9–11^. Recent studies have looked at the association between PRS and breast cancer outcomes. One group developed a breast cancer PRS for risk of recurrence and found that it was associated with worse survival ^12^. The association was comparable to an existing PRS for estrogen receptor negative versus positive risk, however the effect did not appear to be mediated by ER status. Another study found that a breast cancer PRS was associated with less aggressive tumors (lower grade, hormone receptor positive, and smaller tumor size) ^13^. Higher PRS was associated with better OS in unadjusted models; however, the associations were no longer significant after adjusting for clinicopathologic characteristics and treatment.

Here, we investigated the association between cancer and related trait PRS and clinical outcomes in 32,797 (23,770 European) patients treated at the Dana-Farber Cancer Institute. These patients underwent routine tumor sequencing followed by germline imputation using a variety of clinically relevant index dates. This large data set is ideal for interrogating PRS-outcome associations, as it contains both germline and somatic information in addition to clinical information including treatment and outcomes. We conducted a scan of 209 PRS across all cancers as well as within the six most common cancer types in our cohort. We found that a breast cancer risk PRS is associated with improved survival at multiple index dates. This association remained significant when we included multiple relevant covariates and was replicated using public PRS. Interestingly, we found that this association was strongest when restricting analysis to patients with ER-negative tumors. Finally, we identified a similar protective effect of a breast cancer PRS within colon cancer patients receiving palliative-intent therapy, in addition to associations between disease PRS and survival in a pan-cancer analysis.

## Results

### Overview of data

32,797 patients were biopsied as part of routine clinical care at DFCI. Patients were restricted to those most similar to 1000 Genomes European samples in Principal Component Analysis (PCA; see Methods), leaving 23,770 samples. 209 PRS, covering 37 cancer/cancer risk PRS and 63 autoimmune/immune-related PRS (see Methods for PRS details), were evaluated for association with overall survival within six common cancer types: non-small cell lung cancer (NSCLC), colorectal cancer, breast carcinoma, glioma, ovarian cancer, and prostate cancer. This selection of PRS was intended to cover cancer risk, immune-related risk, non-cancer medical conditions, and “control” PRS which were not expected to be associated with survival. Three clinically relevant index dates were investigated: sequencing date, available for all patients; diagnosis date, available for 53% of patients; and first palliative intent therapy, available only for the 25% who received palliative intent treatment at DFCI (Figure 1). Only patients for whom we had the relevant index date were included in each individual model.

**Figure 1.**
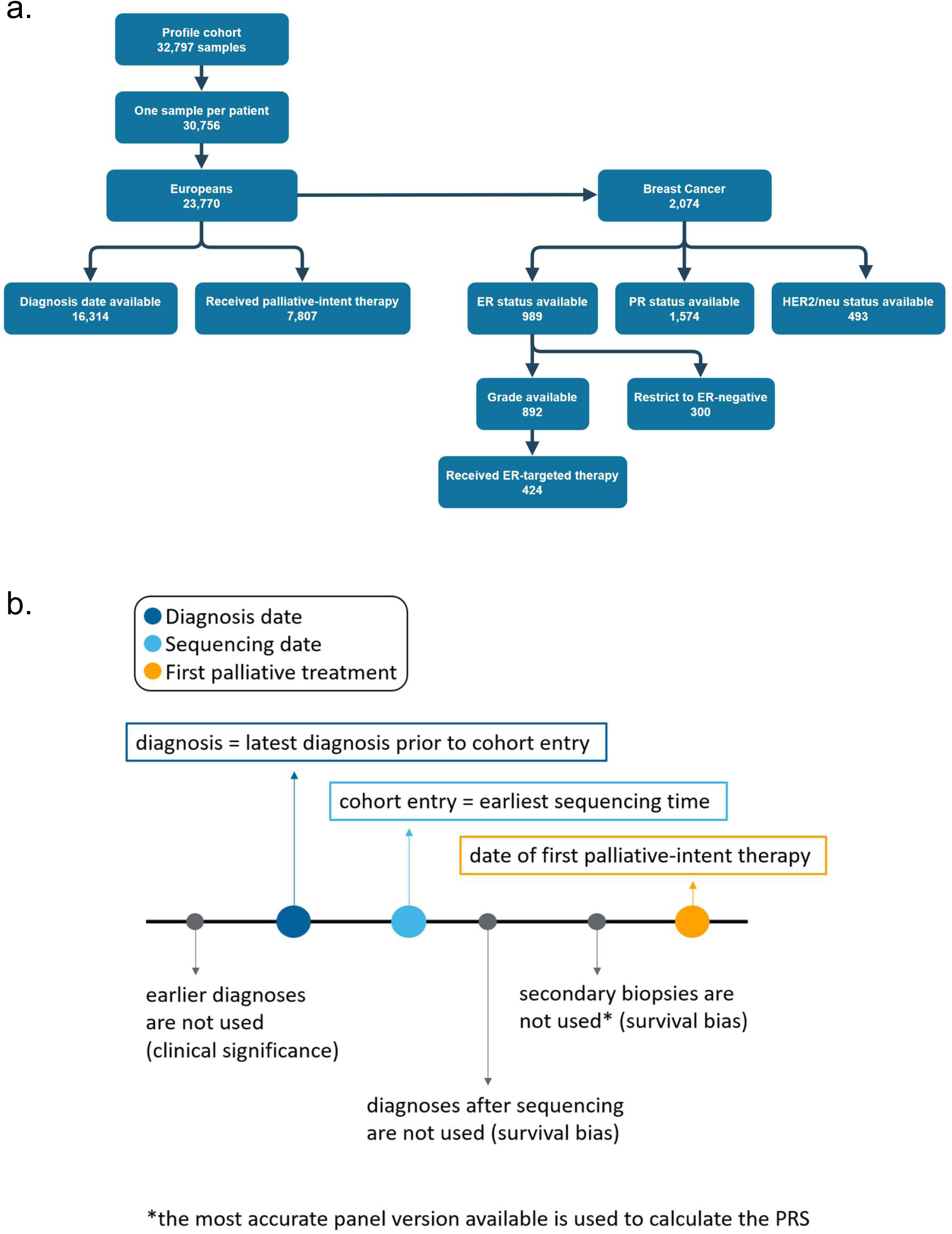
Study cohort and time definitions. a) Breakdown of sample counts in this study. All numbers reflect unique patient counts (1 sample per patient) unless otherwise specified. b) Survival analyses were performed using multiple index dates. The earliest sequencing date was used as the time of cohort entry, and the latest diagnosis before cohort entry was used as the diagnosis in the cohort. Earlier diagnoses and diagnoses after sequencing date were not used in the model. In patients who had multiple biopsies of the same tumor, only the first sample was included in the evaluation. As sequencing panels have varying levels of accuracy, the best sequencing panel was used to generate the PRS.

A Cox Proportional-Hazards survival analysis was performed. This model evaluates the association between the survival time of patients and one or more variables, with respect to known risk factors (covariates). For each patient, covariates were included for age at time of sequencing, sex (where appropriate), version of sequencing panel used, metastatic status of the sample tested, Ashkenazi Jewish ancestry, and five principal components corresponding to genetic ancestry. Tumor stage, ascertained at the time of diagnosis, was included in the models that used diagnosis date as an index date. Stage information was available for 63% of patients in the final palliative treatment cohort and 49% of patients included in the final sequencing index date cohort. Stage information was therefore excluded in the palliative and sequencing index date cohorts. In the pan-cancer analyses, cancer type was included as a categorical covariate with any cancer types having fewer than 500 cases grouped into a separate “Other” category. Patients were excluded who had death or loss to follow up before or the same day as the index date.

### Cancer risk PRS are elevated in the relevant cancer types

We first confirmed the association between cancer risk PRS and the corresponding cancer types, using all other cancers as “controls” since our cohort does not contain patients without cancer who could serve as controls. Nine of fifteen cancer types showed a statistically significant association with a corresponding cancer PRS (Table 1). For example, a breast cancer PRS was associated with breast tumors with an OR of 1.52 per PRS SD and a p of 1.10×10^-67^. The largest OR was observed for thyroid cancer (OR = 1.84, p = 2.60×10^-34^) and prostate cancer (OR = 1.75, p = 2.10×10^-39^).

**Table 1.** Strongest PRS associations with cancer risk within cancer types.

Interestingly, four cancer risk factor PRS were associated with cancer type. Three associations had clear implicated risk factors: a PRS for colon polyps was associated with colorectal cancer (OR = 1.40, p = 2.70×10^-50^), a PRS for Barrett’s esophagus was associated with esophagogastric carcinoma (OR = 1.28, p = 3.80×10^-14^), and a PRS for bronchitis frequency was associated with head and neck carcinoma, likely overlapping with smoking risk (OR = 1.22, p = 1.70×10^-5^). Three additional associations had unclear etiology and were borderline significant after multiple testing: a PRS for high blood pressure was associated with renal cell carcinoma (OR = 1.20, p = 2.10×10^-4^), a PRS for deep vein thrombosis had the most significant association with pancreatic cancer (OR = 1.13, p = 1.10×10^-4^), and a PRS for peripheral artery disease was associated with lower risk for glioma (OR = 0.90, p = 2.10×10^-5^; note because other cancers were used as a control this could alternatively be interpreted as high risk for non-glioma cancers.

### Breast cancer PRS is associated with longer survival in breast cancer patients

We tested each PRS for association with survival within each of six common cancers. Survival was defined as the time from the index date to time of event or loss to follow-up (Figure 1). For breast cancer patients, a PRS for ER-positive breast cancer (hereafter referred to as PRS_ER+) was significantly associated with longer survival (HR = 0.89, p = 1.50×10^-4^) and remained significant after FDR and Bonferroni correction at the sequencing index date (Table 2, Supplementary Figure 1). An additional fifteen PRS were nominally significant, including four breast cancer-related PRS which were all associated with longer survival with the sequencing index date (Supplementary Table 1). Two breast cancer-related PRS were nominally significant with the diagnosis index date, including PRS_ER+ (HR = 0.90-0.91, p = 6.49×10^-3^-6.85×10^-3^). PRS_ER+ was also nominally significant when using the first palliative index date (HR = 0.91, p = 8.25×10^-3^). In every instance, higher breast cancer-related PRS was associated with longer survival. Surprisingly, PRS_ER+ was significantly associated with longer survival in colon cancer using the first palliative index date and remained significant after multiple test correction (6% FDR, HR = 0.85, discussed further below). PRS_ER+ was not associated with survival in other cancer types tested.

**Table 2.** Bonferroni-significant associations between PRS and survival at three different index dates (sequencing date, diagnosis date, and date of first palliative-intent therapy). Models excluding TP53, including TP53, and including the interaction between TP53 and PRS were generated. There was insufficient power for most TP53*PRS analysis, but significant results are included. NS = not significant.

We replicated the association using previously published GWAS data ^14^ and the pgsc_calc pipeline ^15–24^. Seven breast cancer-related PRS were calculated for each patient in our cohort using weights from a public GWAS (Supplementary Note 1). We then tested each score for association with survival. Four scores (ER-positive breast carcinoma, Luminal A breast cancer, breast carcinoma, and Luminal B breast carcinoma) were significantly associated with longer survival and remained significant after multiple test correction (Supplementary Table 2). Similar to PRS_ER+, the ER-positive breast cancer PRS was most significantly associated with longer survival (HR = 0.91, p = 2.68×10^-3^).

### Survival association beyond hormone receptor/treatment status

We considered potential mediating factors that could explain the association between PRS_ER+ and survival. First we sought to determine whether ER status was driving this association. When restricting to ER-negative samples (n = 451), PRS_ER+ remained nominally significant with a similar HR (HR = 0.88, p = 2.50×10^-2^) (Table 3). ER positivity was, unsurprisingly, associated with longer survival in breast cancer patients (HR = 0.65, p = 3.20×10^-11^) (Supplementary Table 3); however, PRS_ER+ remained significant with a similar HR when including ER status as a covariate (HR = 0.92, p = 1.40×10^-2^). When restricting to ER-not-positive cases, PRS_ER+ remained significant and associated with longer survival (HR = 0.85, p = 2.80×10^-3^). When restricting to ER-positive case, the association was no longer statistically significant though directionally consistent (HR = 0.938, p = 0.183).

**Table 3.** Hazard ratios for the two most significant PRS at three clinical index dates: sequencing date, diagnosis date, and first date of palliative-intent therapy. Results are shown for models including a variety of covariates. ER, PR, and HER2/neu indicates patients whose tumors were ER, PR, and HER2/neu-positive, respectively. ER-indicates patients whose tumors were ER-negative. NP = ER-not-positive and includes both patients with ER-negative and ER-unknown tumors. TP53 indicates patients who were positive for a TP53 mutation.

Progesterone Receptor (PR) and HER2/neu status was available for 1,574 patients and 493 patients, respectively. Analysis was restricted to patients for whom PR and HER2/neu status were available. PRS_ER+ remained significantly associated with longer survival when including ER, PR, and HER2/neu status as covariates (HR = 0.93, p = 1.50×10^-2^) (Table 3). To assess the effect of treatment status, we ran a survival model limited to breast cancer patients who received ER-targeted therapy (424 patients receiving tamoxifen, abemaciclib, or fulvestrant). The date of first treatment with ER-targeted therapy was used as the index date for the survival analysis. PRS_ER+ was significantly associated with longer survival (HR = 0.87, p = 4.62×10^-2^). We evaluated multiple additional clinical and demographic characteristics, but none substantially changed the results (Supplementary Note 2).

### Breast cancer PRS is associated with somatic TP53 mutations

We reasoned that the PRS_ER+ association may lead to differences in the somatic presentation of breast cancer tumors. Indeed, high PRS_ER+ was significantly associated with fewer TP53 mutations (OR = 0.85, p = 8.70×10^-5^) (Figure 2, Supplementary Tables 4 and 5). TP53 mutations were not associated with any other PRSs nor was PRS_ER+ associated with any other somatic mutation in breast cancer. To determine whether TP53 status mediated longer survival, we included TP53 mutation status as a covariate in our model. PRS_ER+ remained significant with a similar HR when including TP53 status in the model (HR = 0.93, p = 8.85×10^-3^) (Table 2).

**Figure 2.**
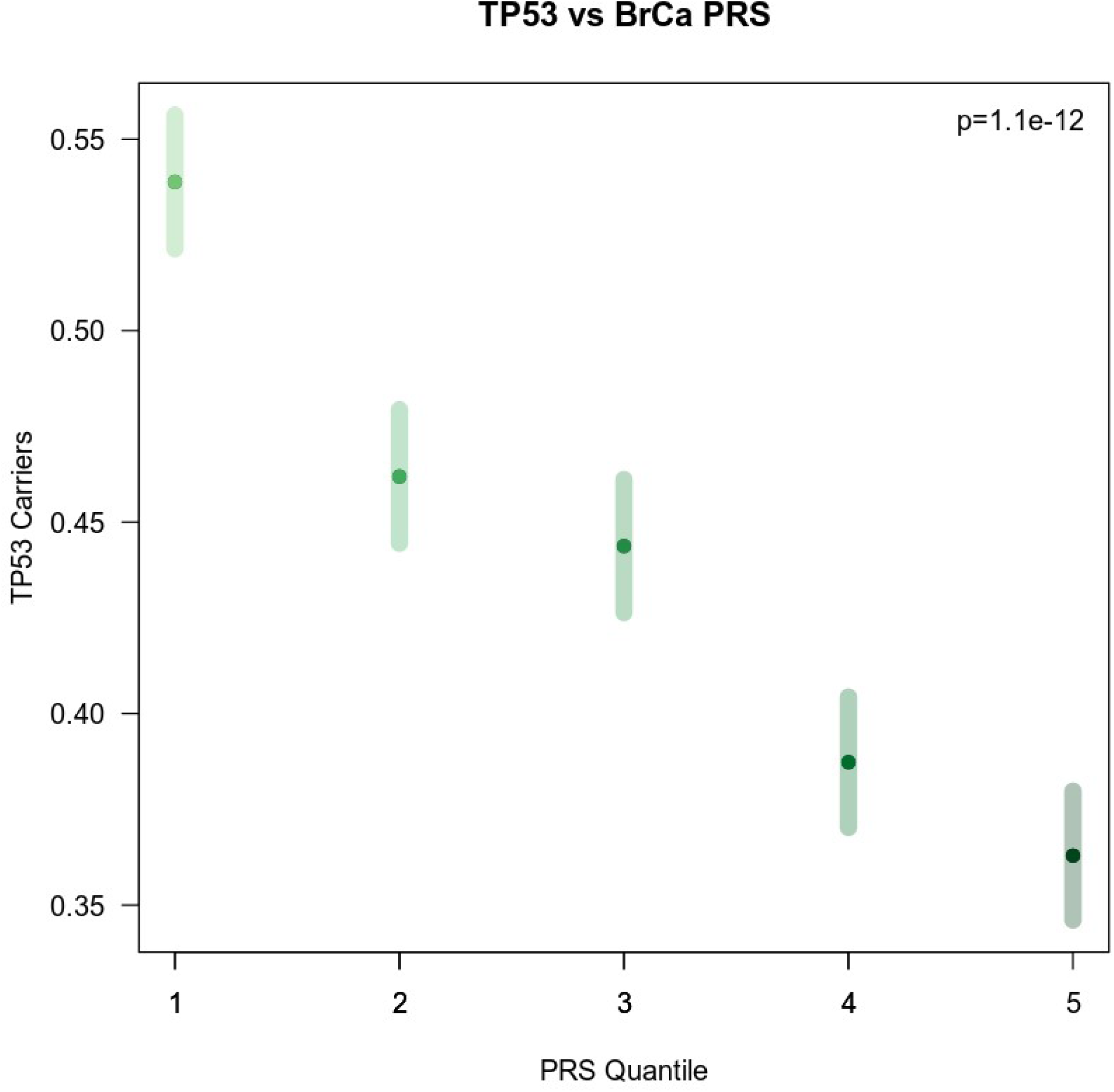
Somatic TP53 mutations versus PRS quartile. Association between somatic TP53 mutations and PRS_ER+. Patients in the highest quartile of polygenic risk demonstrated the fewest TP53 mutations. Patients with the lowest polygenic risk had the highest rate of TP53 mutations.

We hypothesized that the PRS may be associated with the timing of TP53 mutations in mutation carriers. To that end, we used subclonal mutations as a proxy for later developing mutations (defined by low Variant Allele Fraction, see Methods). Indeed, a breast cancer PRS was nominally associated with sub clonal TP53 mutations (OR = 1.22 per PRS SD, p = 0.03) after restricting to mutation carriers, indicating that high-PRS individuals that developed TP53 mutations generally did so later or in subclonal populations. Statistical significance was evaluated by logistic regression with covariates for age, tumor purity, mutation coverage, somatic amplification, and somatic deletion status. Besides TP53, the breast cancer PRS was associated with three additional somatic features in breast cancer (after Bonferroni correction for the number of mutations tested in breast cancer): BRCA1 SNVs, PIK3CA SNVs, and MAP2K4 deep deletions (Supplementary Table 6). After adjusting for TP53 status, the associations with BRCA1 and PIK3CA SNVs became less but still nominally significant. We observed seven additional associations between cancer risk PRS and somatic mutation across the other cancer types, suggesting that germline risk may shape the tumor evolutionary process more broadly (Supplementary Note 3). We estimated the potential influence of variant-level index event bias using the indexevent package ^25^ and did not identify any individual SNPs that appear to be driving the association via index event bias alone (Supplementary Note 4).

### Additional associations

As an exploratory analysis, we tested for associations between PRS_ER+ and survival within five other cancer types to evaluate cancer type-specificity. Surprisingly, in colon cancer patients who received palliative-intent therapy, PRS_ER+ was significantly associated with longer survival (HR = 0.85, p = 2.94×10^-4^) and remained significant after Bonferroni correction (Table 2). This association was not seen with other cancer types or when including patients who never received palliative-intent therapy. The association remained significant when including TP53 status as a covariate. The remaining four breast cancer PRS were also nominally significant with or without including TP53. An additional association was identified between an IBS PRS and longer survival within lung cancer patients when using diagnosis date as the index date.

We then tested every available disease PRS for association with pan-cancer survival across thirteen common cancer types. Bonferroni-significant associations were observed for two type 2 diabetes (HR = 1.04, p = 3.49×10^-4^), bipolar (HR = 1.04, p = 1.84×10^-4^), and pancreatitis (HR = 1.04, p = 2.06×10^-4^) scores using the sequencing index date (Table 2). In all three instances the associations were hazardous. There were no Bonferroni-significant scores with the diagnosis or palliative index dates.

## Discussion

Here we present a large single-institution study of the relationship between germline polygenic risk and cancer outcomes. Higher polygenic risk for ER-positive breast cancer (PRS_ER+) was associated with longer survival and fewer TP53 mutations in breast cancer patients of European ancestry. This association remained significant after adjusting for various clinical and demographic factors. ER-positive breast cancers tend to grow more slowly and generally have a favorable prognosis. However, we found that this association remained significant when limiting to ER-negative tumors and accounting for hormone receptor status. Breast was the only tested cancer in which corresponding cancer risk was associated with survival, indicating that this relationship is not a common phenomenon across all cancer types. However, cancer risk scores were significantly associated with somatic driver status in multiple cancers, implying that germline risk may influence tumor evolution.

PRS have primarily been used to predict an individual’s risk of developing cancer, though recent studies have begun to explore associations with related outcomes. A breast cancer PRS was associated with lower probability of distant metastases and lower probability of breast cancer-related mortality ^13^. However, in women with invasive breast cancer, these associations disappeared after adjusting for treatment and clinicopathologic factors. In contrast, the association we observed in our study remained significant after adjusting for clinicopathologic factors. Notably, lower PRS_ER+ was associated with higher-grade (grade 3) tumor, and higher-grade tumors were associated with shorter survival. By including tumor grade as a covariate in our survival model, we were able to account for this relationship, resulting in a more significant association between germline risk and survival. These findings may indicate clinical context specificity to the association with germline risk.

Breast cancer PRS have been shown to preferentially predict certain cancer subtypes and be associated with TP53 mutations, consistent with our findings ^8^. Beyond breast cancer, European males with higher genetic risk had early diagnosis and fewer driver genes mutated in prostate cancer ^26^. Similar to our findings, higher polygenic risk was associated with better outcomes. However, the association in prostate cancer was largely explained by age. In our analysis, PRS_ER+ remained significantly associated with survival when including age in the model. Moreover, we found an additional association of PRS_ER+ and longer survival in colon cancer patients who received palliative-intent therapy. To date, most studies have found no statistically significant association between cancer risk PRS and either survival or more aggressive cancer ^26–29^. Our PRS associations raise the question of additional PRS-outcome associations in specific treatment settings. Host germline factors have been shown to predict immunotherapy-related toxicities after treatment with immune checkpoint inhibitors ^30^. A PRS for hypothyroidism also predicted the development of thyroid immune-related adverse events in NSCLC patients ^31^. Further studies with longitudinal patient data are needed to investigate treatment specificity.

We consider several potential mechanisms for this association. Prior studies have proposed a polygenic two-hit model ^26^ that can lead to less aggressive tumors. In this model, the patient’s high germline risk serves as the “first hit” for cancer development, thus requiring fewer or less severe somatic alterations for tumorigenesis. This model is consistent with recent observations that rare germline risk variant carriers tend to have fewer somatic drivers ^34^. TP53 is a known early mutation event across cancers and in breast cancer ^35^. Due to the lack of TP53 as a driver in high PRS_ER+ cases, these tumors may be less aggressive overall, resulting in longer survival. The associations between PRS and somatic mutations in breast and other cancers suggest that germline risk may generally impact the acquisition of somatic drivers.

A second possibility is that PRS_ER+ is capturing a spectrum of ER positivity. Some patients with a high PRS_ER+ who develop breast cancer may be expressing ER at a level too low to be detected by immunohistochemistry (IHC). This would be consistent with the observation that PRS_ER+ was still associated with survival in ER-negative patients as well as patients on ER-targeted therapies, where it may be indexing more granular levels of ER-positivity. Sampling error or tumor heterogeneity can lead to false negative ER IHC ^36–38^, which may be captured by the PRS_ER+. Further studies to correlate histology with PRS may help to elucidate the mechanism behind longer survival in patients with high PRS_ER+. Furthermore, digital pathology may prove useful for the detection and quantification of ER positivity, particularly in cases that are ER-negative by IHC.

Finally, we caution that the association may alternatively be explained by an environmental mediator through collider/index event bias. If patients that are at low germline risk need more disruptive environmental events to develop cancer, they may then go on to have poorer outcomes due to those environmental causes. The observed association would still be accurate in a predictive sense, in that high germline risk patients would exhibit longer survival in independent cohorts, but germline genetics would not be the causal factor. This would imply that improvements in breast cancer survival could be achieved with environmental modifications; specifically by avoiding environments observed in low PRS cases. However, we investigated variant-level index event bias in our analysis and did not observe any outliers. We also tested for whether high germline PRS was significantly negatively associated with any other EHR diagnoses, but did not observe any associations that implicated an environmental explanation. Structured collection of additional risk factors and epidemiological measurements, especially in low-PRS cases, may be helpful to identify any additional epidemiological factors that may be driving the PRS-survival association.

Our study had a number of limitations. This single-institution study was restricted to individuals of European ancestry, and it is known that PRS do not translate well across ancestries ^1, 39^. As a cancer institute that receives frequent referrals from other centers, our patient population may consist of more advanced cases. We may be more likely to see patients for whom treatments elsewhere have already failed, which could limit the generalizability of our findings. Moreover, our controls are all patients who have other cancer types, and not patients without cancer. This may also affect the generalizability of our findings. Our study was also limited by the availability of reliable hormone receptor status, tumor stage, and tumor grade information. Hormone receptor status was retrieved from free text in pathology reports, potentially leading to incomplete or inconsistent data. Tumor stage and grade was additionally only recorded for a subset of the patient cohort linked to the cancer registry.

The association between PRS_ER+ and survival may have clinical applications for screening, clinical decision making, and clinical trials. For screening, this association implies that individuals identified as high risk by a PRS are already expected to have longer survival, which may need to be factored into cost/benefit analyses of screening programs ^32^. This may additionally motivate GWAS and development of subtype-specific scores for more aggressive forms of breast cancer ^7^. In the case of clinical decision making, low PRS are indicative of worse survival and may be incorporated into models of patient prognosis. If indeed PRS_ER+ is identifying patients across a spectrum of ER positivity, it may be possible to use PRS_ER+ to prioritize some patients for ER-targeted therapies in the absence of IHC-confirmed ER positivity. Finally, in clinical trials, it may be possible to use PRS to identify a population of patients expected to have longer survival, thus increasing the statistical power of a clinical trial at a fixed budget ^33^. While clinical applications of PRS are still in their early stages, our study highlights how PRS may have continued influence on patients who develop disease. Such PRS may serve as a means of better stratifying patients for screening, informing treatment decisions, matching patients with clinical trials, or providing personalized prognostic information.

## Methods

The Dana-Farber Cancer Institute Profile cohort consists of 32,797 patients biopsied as part of routine clinical care and sequenced on a targeted next-generation sequencing (NGS) panel. Clinical data are available on a substantial portion of patients in the cohort, and new patients are being added to the cohort daily. For our analysis, samples were limited to individuals of European ancestry (23,770). These data were combined with stage information available through the DFCI Cancer Registry. Due to the nature of the Cancer Registry, cancer diagnosis and tumor stage information was inconsistently available within the cohort, with tumor stage information available for 11,205 patients in the cohort. All participants provided informed consent for research. Cohort characteristics are listed in Supplementary Table 7.

### Generating the PRS

We evaluated 209 PRS through collaboration with 23andMe, Inc.. PRS were computed for each patient for common traits including cancer risk and non-cancer, autoimmune disease, cardiovascular disease, and metabolic phenotypes. All PRS were trained in individuals of European ancestry. PRS details are listed in Supplementary Table 8. 206 of the PRSs were trained from data from the 23andMe, Inc. Research Cohort ^40^. Individuals included were research participants of 23andMe, Inc., a direct-to-consumer genetics company, who were genotyped as part of the 23andMe Personal Genome Service. Participants provided informed consent and volunteered to participate in the research online, under a protocol approved by the external AAHRPP-accredited IRB, Ethical & Independent (E&I) Review Services. As of 2022, E&I Review Services is part of Salus IRB (https://www.versiticlinicaltrials.org/salusirb). Five of the 23andMe PRS are denoted genotyped indicating the underlying PRS models are trained on the genotype array data, and are based on an individual-level data model trained using a penalized logistic regression ^41^. The remaining 201 23andMe PRS are denoted imputed as the underlying PRS models are trained on imputed data using a stacked pruning and thresholding approach ^42^. All 23andMe case/control PRS included had an AUROC ≥ 0.55 in a held out test set consisting of individuals of European ancestry. The other 3 PRS are from the PGSCatalog from a previous publication constructed on the UK Biobank cohort for autoimmune traits ^43^. 88 of the PRS are categorized as “Control” PRS which were not intended to have associations with any of the outcomes. Some PRS descriptions end in ‘dx_or_fh’ meaning they were derived from genome-wide association studies where cases had the condition or had family history of the condition.

### Defining clinically relevant index dates

Models were developed using three clinically relevant index dates (Figure 1). Sequencing-based cohort entry was defined as the earliest sequencing date of the patient. Diagnosis-based cohort entry was defined as the date of the last diagnosis prior to cohort entry. Treatment-based cohort entry was defined as the date of the patient’s first palliative-intent treatment in the EHR. Any secondary biopsies after sequencing were not used in the model. For diagnosis-based and treatment-based entry points, potential immortal time bias was accounted for by left truncation until sequencing (at which point the participants formally enter the cohort and survival is no longer biased). Diagnoses earlier than the most recent diagnosis date before sequencing were also not used, in order to capture the temporally closest diagnosis to the date of sequencing, which is the most clinically relevant diagnosis. Death from any cause was defined as our endpoint or “event”, ascertained from DFCI clinical records. Survival was defined as the time from the index date to time of event or loss to follow-up. In our cohort, cancer was the cause of death in approximately 74% of patients.

### Cox proportional hazards analysis

We ran a Cox-proportional hazards analysis, stratified by cancer type, using the R survival package ^44^. Multiple test correction was performed with Bonferroni and FDR methods. Covariates included in the model varied according to index date and tumor type. All analyses included patient age at sequencing, NGS panel version, and genetic ancestry. Metastatic status, patient sex, and tumor stage were also included as appropriate. Pan-cancer and within cancer type analyses were performed for three clinically relevant index dates (Figure 1, Supplementary Notes 2 and 5).

Due to the low number of male patients within the breast cancer subset (n = 16), only females were included in the breast cancer subset to avoid biasing the model. The patient’s sex was therefore not included as a covariate in the breast cancer model. Similarly, the vast majority of glioma cases were listed as primary tumors, with only three being labeled as metastatic within the data set. We therefore excluded all samples labeled as metastatic from the glioma analysis and removed metastatic status from the covariates in that model.

Stage information was not available for all patients in our cohort. To keep the highest number of patients in our models, we grouped patients by tumor stage (1-4), and any patients for whom tumor stage was unavailable was put into a fifth “unknown” group. Including stage information did not improve the predictive power of our models, therefore stage was not included as a covariate with sequencing or treatment index dates. As we did have stage information on patients for whom we had diagnosis date information, stage was included as a covariate in the diagnosis index date analysis. Additional clinical and demographic features were evaluated as detailed in Supplementary Notes.

## Data Availability

23andMe GWAS summary statistics used to generate PRS for this study are not publicly available outside of a collaboration context. Public weights used for this study are available in the PGS Catalog (https://www.pgscatalog.org/).

## Figure Legends

**Supplementary Figure 1.**
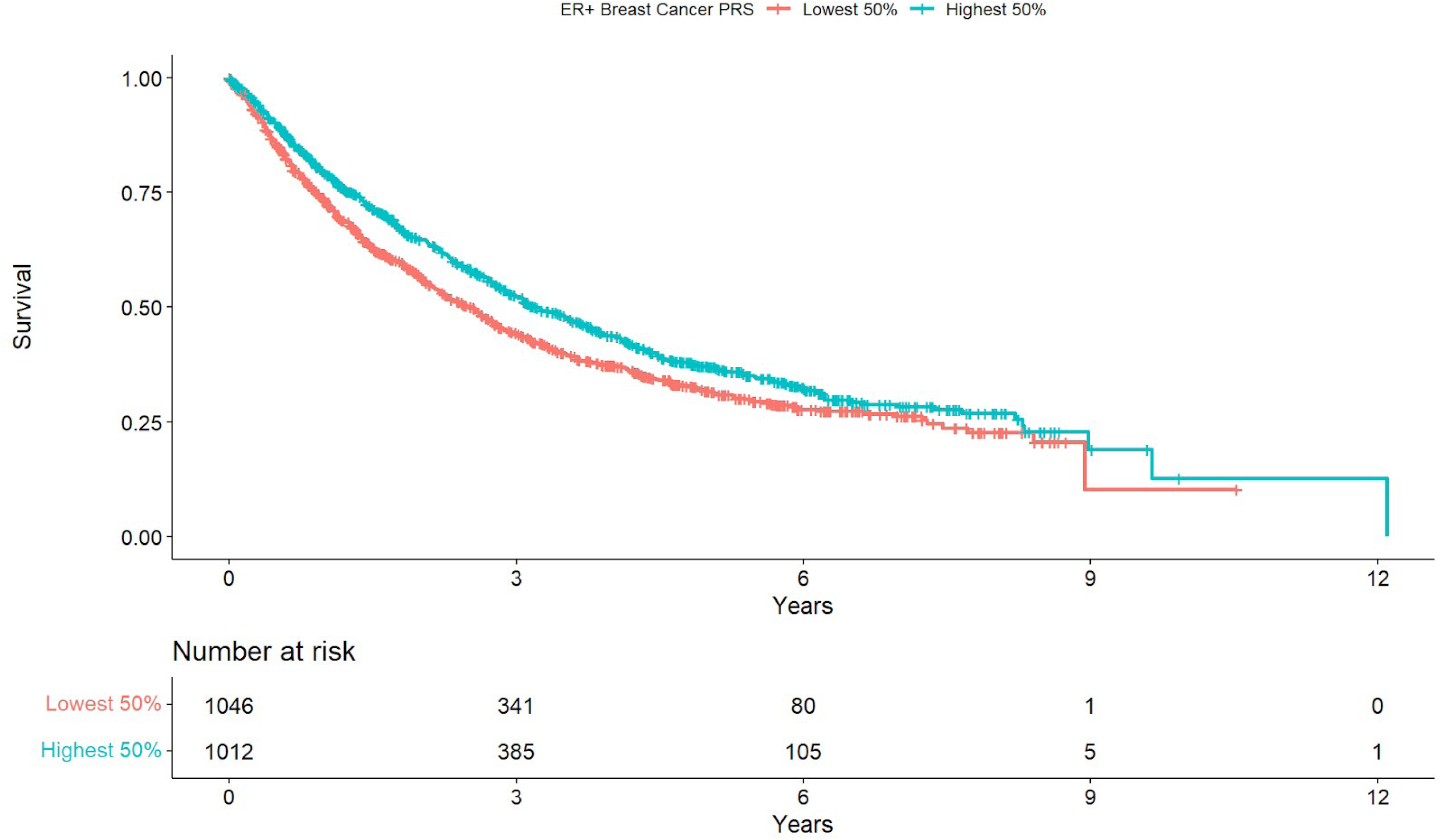
Kaplan-Meier survival curve for PRS_ER+. Kaplan-Meier plot comparing survival of patients with PRS_ER+ scores in the highest 50% versus the lowest 50% of the cohort using the sequencing index date.

**Supplementary Figure 2.**
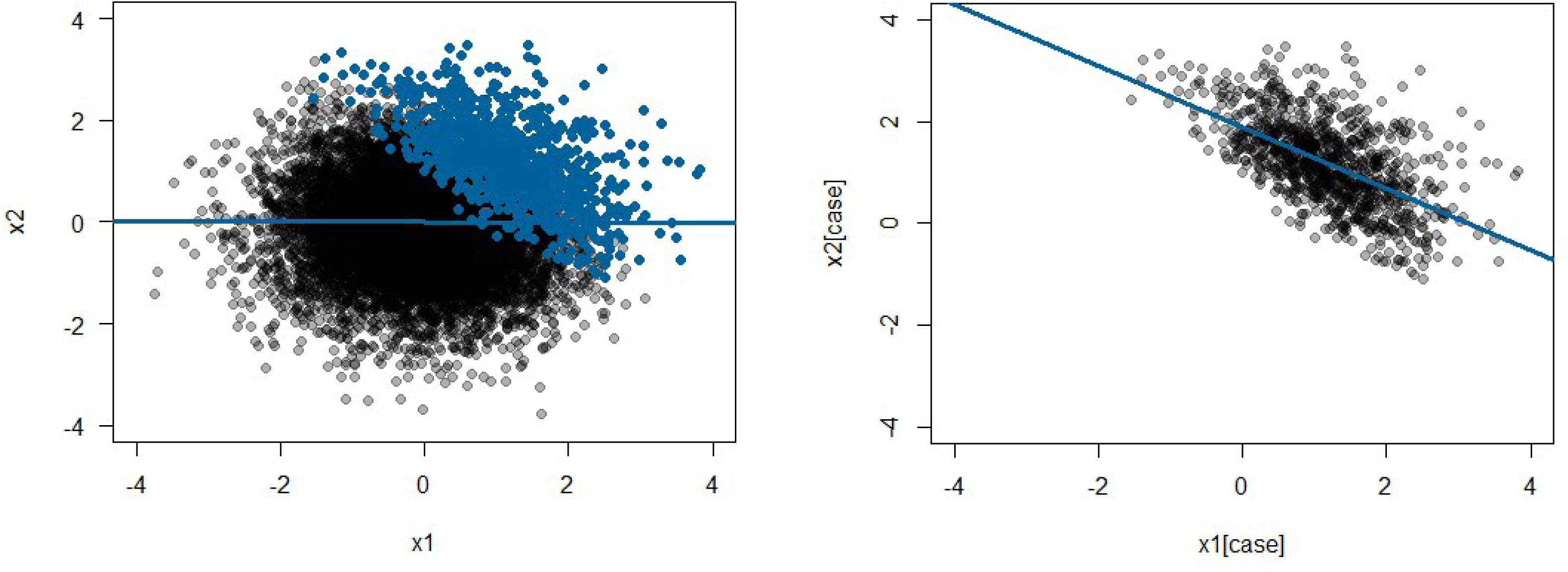
Simulated ascertainment bias. An example distribution of patients is shown on the left, with the top decile highlighted in blue. When we plot a regression on the entire population, no association is seen. However, when we limit our analysis to the top decile of patients, we see an association.

**Supplementary Figure 3.**
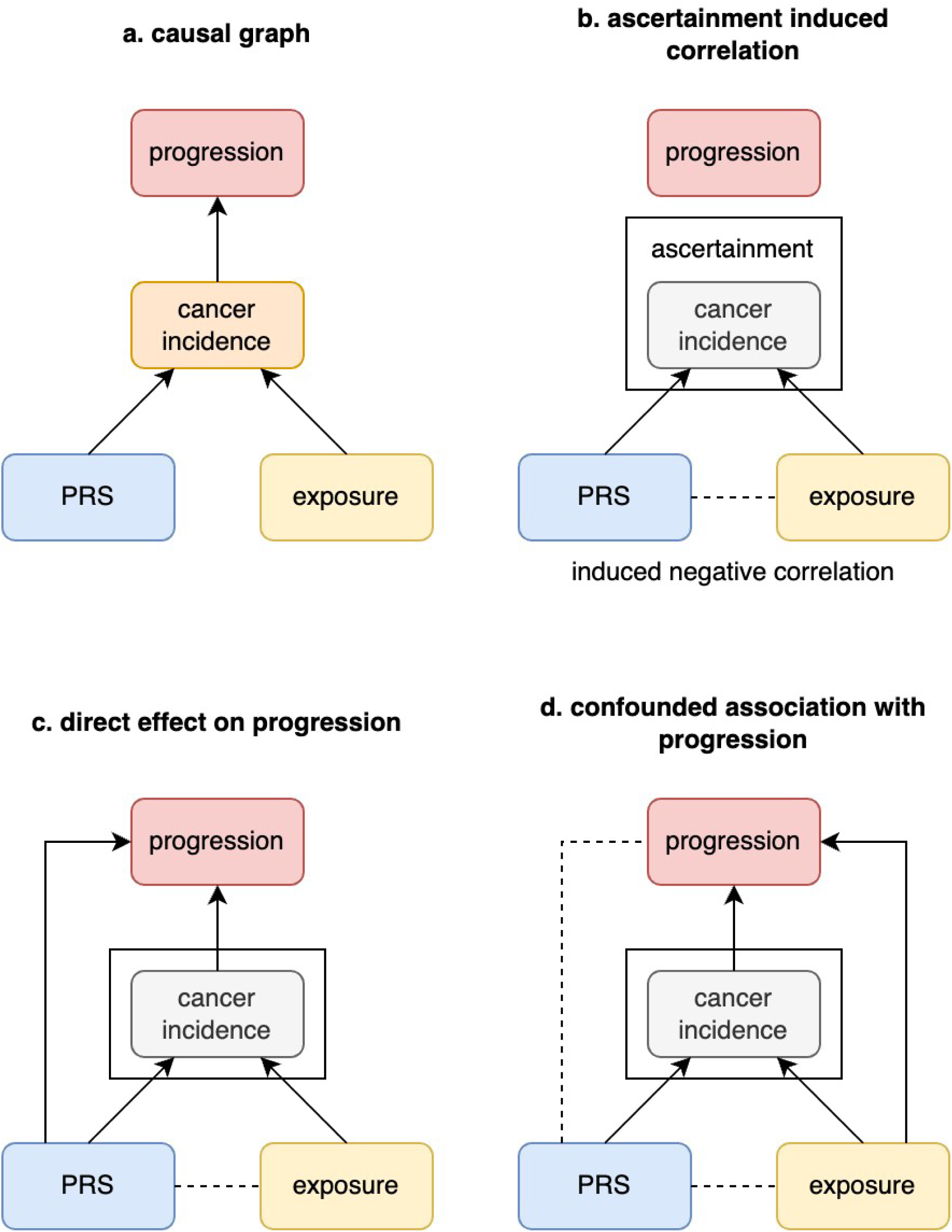
Relationships between genetic risk and exposure on cancer progression. a) Both genetic risk (PRS) and environmental exposures contribute to cancer incidence. b) When we ascertain a cohort based on cancer incidence, we may identify an induced negative correlation between genetic risk and environmental exposure. c) Genetic risk may contribute to cancer progression independently of environmental exposure. This may appear to show a negative correlation between genetic risk and environmental exposure. d) Where environmental exposure contributes independently to progression, we may see an induced negative correlation between genetic risk and progression, and genetic risk and exposure.

**Supplementary Figure 4.**
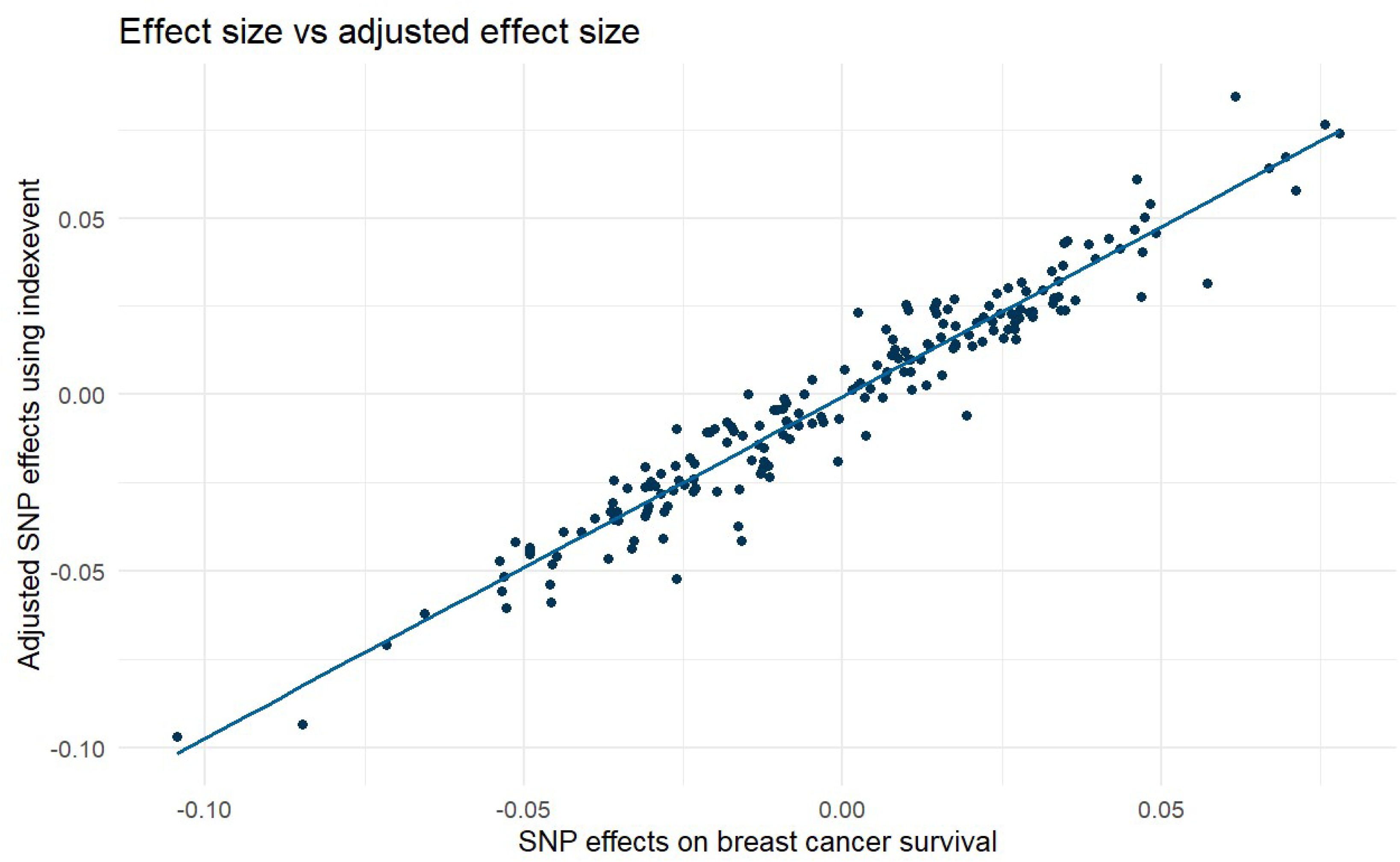
Comparing raw and bias-adjusted SNP effects. Comparison between raw vs corrected SNP effects on breast cancer survival using the indexevent package in R.

## Table Legends

**Supplementary Table 1.** All tested associations between PRS and survival at three different index dates (sequencing date, diagnosis date, and date of first palliative-intent therapy). Models excluding TP53, including TP53, and including the interaction between TP53 and PRS were generated. There was insufficient power for most TP53*PRS analysis, but significant results are included.

**Supplementary Table 2.** Survival associations between PRS and survival using the sequencing date index date. PRS were calculated for breast cancer patients within the Profile cohort from public GWAS weights obtained via the PGS Catalog.

**Supplementary Table 3.** Association between ER status and survival within breast cancer patients at three clinical index dates.

**Supplementary Table 4.** The most significant associations between somatic mutations and cancer-related PRS within cancer types.

**Supplementary Table 5.** All Bonferroni-significant associations between breast cancer PRS and Tier 3 TP53 mutations in breast cancer.

**Supplementary Table 6.** All Bonferroni-significant associations between PRS_ER+ and somatic alterations in breast cancer.

**Supplementary Table 7.** Characteristics of the study cohort. Study was limited to Europeans (n = 23,770). After calculating time intervals for survival analyses, samples were restricted to tumors with sequencing dates prior to date of death or loss to followup (n = 22,229). Cancer types with fewer than 500 samples were grouped into "Other". Hormone receptor status was evaluated for all breast cancer patients with an available pathology note (n = 1,673).

**Supplementary Table 8.** Names and phenotypes of all 209 tested PRS. dx = diagnosis. fh = family history.

**Supplementary Table 9.** All significant association between PRS and cancer risk within cancer types. The most significant associations within each cancer type are highlighted.

**Supplementary Table 10.** Association between PRS and grade within breast cancer patients and ER-negative breast cancer patients.

**Supplementary Table 11.** Association between PRS and survival within breast cancer patients using the sequencing index date and including grade as a covariate.

**Supplementary Table 12.** Hazard ratios for all five tested breast cancer PRS. HRs were calculated excluding ancestry, including Ashkenazi Jewish ancestry but excluding principal components 1-5, and including principal components 1-5 but excluding Ashkenazi Jewish ancestry. PC = principal components, AJ = Ashkenazi Jewish ancestry.

**Supplementary Table 13.** CoxPH coefficient, HR, standard error, and p-values of the most significant cancer risk PRS. Models were run within cancer type.

## Acknowledgements

We would like to thank the research participants and employees of 23andMe for making this work possible.

The following members of the 23andMe Research Team contributed to this study: Stella Aslibekyan, Adam Auton, Elizabeth Babalola, Robert K. Bell, Jessica Bielenberg, Ninad S. Chaudhary, Zayn Cochinwala, Sayantan Das, Emily DelloRusso, Payam Dibaeinia, Sarah L. Elson, Nicholas Eriksson, Chris Eijsbouts, Teresa Filshtein, Pierre Fontanillas, Davide Foletti, Will Freyman, Zach Fuller, Julie M. Granka, Chris German, Éadaoin Harney, Alejandro Hernandez, Barry Hicks, David A. Hinds, M. Reza Jabalameli, Ethan M. Jewett, Yunxuan Jiang, Sotiris Karagounis, Lucy Kaufmann, Matt Kmiecik, Katelyn Kukar, Alan Kwong, Keng-Han Lin, Yanyu Liang, Bianca A. Llamas, Aly Khan, Steven J. Micheletti, Matthew H. McIntyre, Meghan E. Moreno, Priyanka Nandakumar, Dominique T. Nguyen, Jared O’Connell, Steve Pitts, G. David Poznik, Alexandra Reynoso, Shubham Saini, Morgan Schumacher, Leah Selcer, Anjali J. Shastri, Jingchunzi Shi, Suyash Shringarpure, Keaton Stagaman, Teague Sterling, Qiaojuan Jane Su, Joyce Y. Tung, Susana A. Tat, Vinh Tran, Xin Wang, Wei Wang, Catherine H. Weldon, Amy L. Williams, Peter Wilton.

We would also like to thank the members of the Gusev lab for their input and helpful conversations.

