## Supplementary Notes for "Association between polygenic risk and survival in breast cancer patients"

**Supplementary Note 1: Replication using public GWAS**

Weights were retrieved from publicly published GWAS data ^1^ from the PGS Catalog using the pgsc_calc pipeline ^2–11^. Seven breast cancer-related PRS were calculated for each patient in our cohort using weights from public GWAS. We then tested each of these for association with survival within the Profile breast cancer cohort with age, NGS panel version, metastatic status, and ancestry as covariates. Note that two of these PRS (PGS000004 and PGS000005) were developed in another study ^12^; however, all seven PRS from the public GWAS were included in our analysis. We confirmed association between the public ER-positive breast cancer PRS and breast cancer (Supplementary Table 9).

**Supplementary Note 2: Evaluating additional clinical and demographic features**

*Age*

As environmental factors can also play a role in breast cancer survival, it was essential to evaluate survival in light of known potential confounders. Age at diagnosis was included as a covariate in the primary model and removing it did not alter the association (p = 1.46x10^-4^ when excluding age from the model). Adding stage at diagnosis as a categorical covariate for the diagnosis index date likewise did not substantially change the PRS association (p = 4.83x10^-4^, HR = 0.90).

*Grade*

Tumor grade was extracted from the EHR. Structured grade information was available for 892 breast cancer patients (Figure 1). All patients without grade 1-4 recorded were batched into a fifth “unknown” category. Cox-proportional hazards analysis was performed within breast cancer patients including ER status, age at diagnosis, panel version, metastatic status, PC 1-5, and tumor grade as covariates. Survival models were generated for both PRS_ER+ and a general breast cancer PRS. Analysis was also performed after restricting to ER-negative patients.

PRS_ER+ was inversely associated with grade 3 tumors (p = 2.58x10^-3^) in the breast cancer cohort; however, when restricting to ER-negative tumors, this association was no longer present (Supplementary Table 10). PRS_ER+ remained significant (HR = 0.93, p = 1.58x10^-2^) when including tumor grade in the model (Supplementary Table 11). Interestingly, restricting to ER-negative patients markedly increased the protective effect seen with high PRS (HR = 0.80, p = 1.91x10^-3^). A generic breast cancer PRS also remained protective when accounting for tumor grade (HR = 0.93, p = 2.43x10^-2^) and demonstrated a substantial increase in protective effect when restricting to ER-negative tumors (HR = 0.78, p = 1.69x10^-4^).

*ER, PR, HER2/neu analysis*

Hormone receptor status was extracted from free-text pathology reports using str_detect from the stringr package ^13^. We first restricted our patient cohort to breast cancer patients. Whether the patients were reported ER-positive, ER-negative, or other (unknown, indeterminate, etc.) was then recorded in a table. PR and HER2/neu status were also extracted from free-text reports in the same manner and recorded for all patients.

*ER-targeted therapy analysis*

Patients who received treatment with tamoxifen, abemaciclib, or fulvestrant were extracted from a DFCI treatment plan database. The first date of treatment with any of these medications was set as the index date for ER-targeted therapy analysis. A Cox-proportional hazards analysis was performed within treated breast cancer patients (n = 424) including age, NGS panel version, genetic ancestry, and metastatic status as covariates. Associations with PRS_ER+ and a general breast cancer PRS were tested.

*Ashkenazi Jewish ancestry*

To compute Ashkenazi Jewish (AJ) ancestry, all European ancestry samples were projected into the corresponding principal component coordinates, as previously defined in a large AJ reference panel. ^14^ As a positive control, AJ ancestry was confirmed to be significantly positively associated with self-reported Jewish religion from self-reported patient demographics in the EHR. Survival analysis was performed including AJ ancestry status as a continuous covariate at all index dates within the breast cancer subset. We performed analyses including the first five principal components (PC 1-5) and excluding AJ ancestry score as a baseline. We then performed survival analysis replacing principal components with the AJ ancestry score. Finally, we performed analyses including both principal components and AJ ancestry as covariates in the models. Inclusion of AJ ancestry as a covariate did not meaningfully change the results: all five breast cancer PRS remained significant for the sequencing date and diagnosis date analysis, and four out of five were significant for the palliative therapy analysis (Supplementary Table 12). In all cases, higher PRS were associated with longer survival at all index dates with or without including AJ ancestry as a covariate.

**Supplementary Note 3: Pan-cancer scan for PRS-somatic associations**

We observed seven additional associations between cancer risk PRS and somatic mutation across the other cancer types after within-cancer Bonferroni correction (Supplementary Table 4). For example, the PRS for lung cancer diagnosis or family history was associated with increased incidence of KRAS mutations in NSCLC patients (OR = 1.17, p = 9.20x10^-6^). This is consistent with prior findings that lung cancer in smokers is primarily KRAS-driven and the lung cancer PRS is highly correlated with smoking. The PRS for melanoma diagnosis or family history was associated with increased incidence of ARID2 mutations in melanoma patients (OR = 1.42, p = 1.20x10^-4^). ARID2 mutations are frequently observed in early melanoma lesions and may thus be indicative of initiating driver events ^15,16^. The PRS for Barrett's esophagus was associated with decreased incidence of CDH1 mutations in esophagogastric cancer patients (OR = 0.64, p =  4.6x10^-5^). CDH1 mutations are elevated in diffuse gastric cancer and may thus indicate Barret’s esophagus as a risk factor for non-diffuse GC ^17^. These findings suggest that germline risk may shape the tumor evolutionary process more broadly, either increasing or decreasing the rate of certain drivers, depending on the cancer. In cancer types where the PRS captures risk for environmental factors, we expect to see a higher frequency of driver mutations; conversely, we would expect to see fewer driver mutations in cancer types for which the risk PRS are not capturing extrinsic risk factors.

**Supplementary Note 4: Index event bias**

By focusing our analysis on patients who developed cancer, we introduce the possibility of collider or “index event” bias driven by the ascertainment. Specifically, if breast cancer is caused by the additive effects of PRS and independent environmental factors, then those factors may become (negatively) correlated in ascertained breast cancer cases (Supplementary Figure 2, Supplementary Figure 3). The association of genetic risk and survival may thus potentially be explained by the direct effect of some unmeasured *environmental* factor that has become negatively correlated with risk due to the ascertainment. To investigate this possibility, we estimated the potential influence of variant-level index event bias using the indexevent package ^18^. This method attempts to adjust for potential confounding effects by incorporating the GWAS effects for the index event itself (in this case cancer incidence) under the assumption that confounding is constant across SNPs and with adjustment for attenuation in the bias estimator. We note that this adjustment specifically assumes that there is no genetic correlation between incidence and prognosis, whereas such a correlation is one possible interpretation of the PRS-survival association we observed. Therefore, our goal with this analysis was to investigate whether individual variants are having an outsized/outlier effect.

The regression-based estimator of bias can be attenuated by estimation error, and two methods for bias correction were proposed: a Hedges-Olkin adjustment, which is a simple analytical procedure that uses the observed variance and standard errors of effects to remove error; and Simulation extrapolation (SIMEX), a simulation-based technique where measurement error is added in increments to data, estimates are computed, and a trend is then extrapolated back to a case with no measurement error ^18–20^. We used Hedges-Olkin and SIMEX adjustments available in the indexevent package in R ^18^. We used the previously published GWAS summary statistics ^1^ for effect sizes and standard errors of the index trait (breast cancer) and the corresponding individual SNP effect sizes and standard errors for the subsequent trait (survival). We observed a significant positive relationship between incidence and prognosis across variants (Hedges-Olkin adjustment coefficient of 0.07 (95% CI: 0.07-0.07), SIMEX adjustment coefficient of 0.11 (95% CI: 0.11-0.11); see Methods). However, plotting the SNP effect size on survival before and after adjustment showed no substantial difference in the overall SNP effect sizes (Supplementary Figure 4). Thus, we did not identify any individual SNPs that appear to be driving the association via index event bias alone.

**Supplementary Note 5: Additional details on clinically relevant index dates**

*Sequencing index date analysis*

Pan-cancer analysis was performed on all samples. Additionally, samples were evaluated within cancer type for cancer types with >1000 unique patients. This included NSCLC (3,185), CRC (2,129), BRCA (2,074), Glioma (1,809), and OVCA (1,142). PRCA (615) was also included due to genetic relatedness to breast cancer, despite having fewer than 1000 samples available.

*Diagnosis index date analysis*

Pan-cancer analysis was performed on all samples. Samples were evaluated within cancer type for cancer types with >1000 unique patients. These included NSCLC (2,552), breast carcinoma (1,508), colorectal cancer (1,499), and glioma (1,165). Prostate cancer was also included in the analysis due to its genetic relatedness to breast cancer, despite having only 527 cases.

*Palliative treatment index date analysis*

Pan-cancer analysis was performed on all samples that received palliative-intent therapy. Due to the relatively small number of the cohort receiving palliative-intent therapy, analysis was performed within cancer type on tumor types with >500 samples. This included NSCLC (1,253), breast carcinoma (1,089), colorectal cancer (729), and ovarian cancer (504). Again, prostate cancer (201) was included due to its genetic relatedness to BRCA, though it did not meet the threshold of 500 patients.

*Additional findings of interest*

Higher PRS_ER+ was strongly statistically associated with longer survival in colorectal cancer patients who received palliative-intent therapy. This association was not observed in other (non-breast) cancer types nor in colorectal cancer when evaluating from diagnosis or sequencing index dates, but was specific to the group receiving palliative treatment. Estrogen-mediated signaling has been hypothesized to be protective in colorectal cancer, with ERβ expression decreasing during tumor progression ^21–25^. We attempted to assess whether this increased survival was due to ER-targeted therapy in these patients; however, only 13 colorectal patients in our cohort had documented ER testing and received targeted ER therapy, which was not sufficiently powered to evaluate treatment-specific survival.
